# Differences in disease characteristics and outcomes as determined by biological sex in a large UK IPF population: Analysis from the British Thoracic Society, Interstitial Lung Disease (BTS-ILD) registry data

**DOI:** 10.1101/2025.03.18.25323768

**Authors:** S. Mulholland, G. Dixon, M Wells, P. White, S. Harding, AM. Russell, SL. Barratt

## Abstract

**Introduction:** Growing evidence suggests that biological sex influences the incidence, presentation, diagnosis and outcomes of many lung diseases. Understanding these differences is the first step towards precision medicine to improve patient care.

**Methods:** In this cross-sectional study IPF patients enrolled in a national, multicentre registry (UK BTS-ILD) were categorised by sex and analysed for differences in demographics, pulmonary function tests, HRCT radiological pattern, eligibility/uptake of antifibrotics and survival.

**Results:** Of 7177 cases, 77.8% (n=5587) were male, median age 75 years (IQR 69.5-80.5) for both sexes (p=0.83). Males were more likely to have a history of smoking (p<0.001) and lower baseline median FVC % predicted (males 76.4%, IQR 66.2-86.7 vs females 78.8%, IQR 68.6-89.1, p<0.001). Diabetes, cardiovascular disease and gastro-oesophageal reflux disease were statistically more common in males (p<0.001), whilst major depressive illness was more common in females (p<0.001). Significantly more females experienced symptoms for >24 months prior to first clinic appointment (females 40.1% vs males 36.6%, p=0.028). Whilst more males in the cohort met eligibility criteria for antifibrotics at baseline (p<0.001), a larger proportion chose not to commence antifibrotic treatment (males 47.0% vs females 29.6%, p<0.001). Female sex was associated with longer mean survival (female 9.99 years (95% CI 9.18-10.79) vs males 8.57 years (95% CI 8.15–8.99), p<0.001). Male sex, higher age, lower baseline FVC % predicted and co-existent lung cancer were all independently associated with worse survival (p<0.001).

**Conclusion:** This is the first UK study to use national registry data to systematically evaluate IPF disease characteristics stratifying by biological sex and highlights distinct characteristics between groups. Future clinical trials should explicitly explore sex-specific targeted interventions and analyses, to optimise future IPF patient care.

**What is already known about this topic?:** Idiopathic pulmonary fibrosis (IPF) is a progressive fibrotic lung condition with a poor prognosis. There are established sex differences in the incidence and prevalence of IPF, and reports of differing lived experience.

**What this study adds?:** This study is the first to report on UK real-world registry dataset to systematically evaluate sex differences in clinical disease characteristics, treatments and outcomes. The results of this study suggest that IPF has distinct characteristics depending on biological sex.

**How this study might affect research, practice or policy?:** These findings highlight key areas that could be addressed to improve patient outcomes and care within the UK. It creates the opportunity to explore tailored approaches to care.

## Introduction

Pulmonary Fibrosis (PF) describes a group of lung diseases where scar tissue progressively and irreversibly replaces normal lung tissue^1^. There are established sex differences in the incidence and prevalence of various subtypes of PF^2, 3^. Idiopathic pulmonary fibrosis (IPF), the archetypal progressive phenotype, is twice as common in males than females^1, 4^. Lived experience of PF differs between males and females. Independent of age, disease severity or functional capacity, females have greater mental health impairment but fewer physical health related QoL impairments than males^2^. The underlying reasons for these variances are not known but small cohorts have implicated differences in environmental exposures^5^and sex hormones^6^ as potential contributing factors. Understanding these differences is the first step towards precision medicine for patients based on biological sex, to improve patient care.

Antifibrotic medications are known to have similar efficacy and adverse event profiles in males and females^7, 8^. Females are significantly underrepresented in these clinical trials. Limited observational registry data currently available has shown males have a shorter time to treatment initiation than females^9^, which cannot be explained by disease severity alone. The impact of physician-patient sex concordance and implicit bias as a factor contributing to healthcare disparities remain unknown in PF.

Systematic evaluation of sex differences in clinical disease characteristics, treatments and outcomes, are urgently needed. We characterised a large UK IPF population by biological sex, determining differences in disease characteristics and outcomes using the British Thoracic Society (BTS) IPF registry data from Jan 2013-October 2024. Baseline characteristics and trends in management of the registry IPF cohort between 2013-2019 and 2013-2023 were previously published^10, 11^, but did not include detailed exploration of sex differences.

## Methods

### Registry platform and data management

The UK ILD (Interstitial lung disease) registry has ethical approval (National Research Ethics Service (NRES) reference 12/EE/0381 and renewal 12/EE/0346) to enrol all patients with a definite or probable IPF diagnosis, in accordance with the American Thoracic Society, European Respiratory Society, Japanese Respiratory Society and Latin American Thoracic Society guidelines (ATS/ERS/JRS/LATS) (Raghu 2011)^12^, providing written consent is obtained. The UK IPF Registry is funded by the British Thoracic Society. Patient and public representatives were involved in the original development of the registry programme and have ongoing representation through patient charities.

Data is collated and entered by the hospital clinical team responsible for the patient’s care, into a secure web-based platform (https://www.brit-thoracic.org.uk/quality-improvement/lung-disease-registries/bts-ild-registry/), that encrypts identifiable data at the point of entry. Data validation occurs at the point of data entry and includes limiting responses within specific data range^12^. Data can be submitted either prospectively or retrospectively (provided first clinic visit was on or after 1^st^ January 2013), at baseline and then at least every 12 months.

### Antifibrotic use within the UK

In England, antifibrotic medications are prescribed by ILD specialist centres, commissioned by the National Health Service (NHS). Antifibrotics are prescribed by general hospitals in Scotland, Wales and Northern Ireland, where commissioning differs. Pirfenidone has been provided through the NHS since 2013 (originally TA282)^13^, with nintedanib approved in 2016 (TA379)^14^. The original technology appraisal guidance for IPF enabled the prescription of pirfenidone or nintedanb for patients diagnosed with IPF who have a Forced Vital Capacity of 50-80% predicted. Recommendations were amended in 2023, additionally authorising the use of nintedanib in patients with an FVC >80% predicted (TA864)^15^. Baseline FVC % predicted became the accepted marker of disease progression to determine the eligibility of patients for antifibrotics.

### Project methodology

A data access request was submitted to the BTS ILD registry (September 2023) and approved by the BTS ILD registry committee in February 2024. Access to the complete dataset from 1^st^ January 2013 (date of origin) to the censor date of 15^th^ October 2024 was obtained in October 2024. Patients with a documented diagnosis of IPF and recorded biological sex were included in the analysis.

Data collated included age, sex, ethnicity, smoking history, duration of symptoms, co-morbidities, presenting radiological pattern of pulmonary fibrosis on HRCT scan, (definite usual interstitial pneumonia(UIP), probable UIP or indeterminate for UIP)^12^, lung function tests (forced expiratory volume in 1 second (FEV1), forced vital capacity (FVC), transfer factor of the lung for carbon monoxide (TLCO) and transfer coefficient of the lung for carbon monoxide (KCO)), 6-minute walk tests (6MWT) and Medical Research Council(MRC) dyspnoea scale, treatments, management and outcomes. GAP stage was calculated (gender, age, physiology) using the available data^16^.

### Data analysis

Data were analysed using SPSS (IBM Corp., Chicago, IL, USA, version 29.0.2.0). Descriptive statistics characterise baseline demographic and clinical characteristics of the patient cohort according to biological sex. In addition, differences were sought between the two sexes on antifibrotic eligibility at baseline (using baseline FVC % predicted as per NHS guidance), the uptake of antifibrotics by patients and the holistic care of patients in terms of oxygen assessment and referral for pulmonary rehabilitation, palliative care or lung transplantation.

Data three standard deviations above or below the median were independently sense checked and excluded from the analysis of that variable if deemed erroneous.

Continuous data were expressed as median with interquartile range and compared using the Mann Whitney U test. Categorical data were expressed as number and percentage and compared using Chi-squared test.

Kaplan–Meier survival curves were constructed to compare biological sex groups and analysed using Log-rank testing; a p-value of less than 0.05 was considered statistically significant. Mortality risk factors were explored using Cox regression analysis among the pre-defined variables of sex, smoking history, baseline FVC and co-morbidities in a subgroup of the cohort with complete datasets and follow up of at least 5 years. Interaction terms were tested to determine whether the effect of covariates differed by sex.

## Results

### Participating hospitals

At the time of data censorship, 64 hospitals were actively participating in the registry (Appendix 1); 13 were specialist ILD centres in England, forty were secondary care centres in England and 11 were centres across the devolved nations (Scotland, Wales and Northern Ireland, where commissioning differs).

### Baseline characteristics

Between 1^st^ January 2013 and 15^th^ October 2024, n=7177 cases of IPF were enrolled into the UK IPF registry with data available.

Baseline demographics are detailed in Table 1. Most patients were male 77.8% (n=5587/7177), with a median age 75 years (IQR 69.5-80.5) for both sexes (p>0.05). Where ethnicity was recorded, White British predominated in both males (n=846, 93.6%) and females (n=254, 92.4%). In February 2023 the question wording changed within the ILD registry to explicitly separate sex and gender questions. Our study is purely concerned with the analysis of sex differences.

**Table 1:** Baseline demographics and characteristics of UK Idiopathic Pulmonary Fibrosis (IPF) cohort between January 2013-October2024, stratified by biological sex. Statistically significant values highlighted in bold. Abbreviations: IQR, interquartile range; n, number; %, percentage; FEV1, forced expiratory volume in 1 second; FVC, forced vital capacity; BMI, Body mass index; m, metres; MRC, (Medical Research Council) dyspnoea scale; GAP, Gender Age Physiology; ILD, interstitial lung disease; p value, statistical probability value.* co-morbidities are not mutually exclusive.

|  | Number of patients with data | Male | Female | P Value |
| --- | --- | --- | --- | --- |
| **Biological Sex (n, %)** | 7177 | 5587 (77.8%) | 1590 (22.0%) | **<0.001** |
| **Age (median, IQR)** | 7102 | 75 (69.5-80.5) | 75 (69.5-80.5) | 0.83 |
| **Smoking Status (n, %)** | 5619 |  |  |  |
| Ever smoker |  | 3229 (72.9%) | 718 (60.5%) | **<0.001** |
| Never smoker |  | 1203 (27.1%) | 469 (39.5%) |  |
| **Ethnicity (n, %)** | 1179 |  |  |  |
| White (British) |  | 846 (93.6%) | 254 (92.4%) | 0.48 |
| Other |  | 58 (6.4%) | 21 (7.6%) |  |
| **Co-morbidities (n, %)\*** |  |  |  |  |
| Cardiovascular disease | 5227 | 2434 (58.9%) | 522 (47.8%) | **<0.001** |
| Diabetes | 5227 | 941 (22.8%) | 169 (15.5%) | **<0.001** |
| Gastro-oesophageal reflux disease | 5227 | 825 (20.0%) | 269 (24.6%) | **<0.001** |
| Major depressive illness | 5227 | 27 (2.5%) | 34 (0.8%) | **<0.001** |
| Lung carcinoma | 5227 | 14 (1.3%) | 42 (1.0%) | 0.45 |
| Other Malignancy | 5227 | 87 (2.1%) | 16 (1.5%) | 0.18 |
| BMI >30 | 3302 | 938 (33.1%) | 285 (35.8%) | 0.16 |
| **Pulmonary hypertension or right heart strain (n,%)** | 907 | 150 (21.4%) | 49 (23.9%) | 0.44 |
| **Baseline lung function (median, IQR)** |  |  |  |  |
| FEV1 % predicted | 5736 | 81.4 (70.5-92.3) | 80.5 (67.5-93.5) | **<0.001** |
| FVC % predicted | 5741 | 76.4 (69.5-80.5) | 78.8 (68.6-89.1) | **<0.001** |
| TLCO % predicted | 4012 | 49.5 (38.7-60.3) | 49.2 (38.9-59.6) | 0.44 |
| KCO% predicted | 3837 | 79.2 (64.8-93.7) | 77.4 (65.5-89.4) | 0.18 |
| **MRC dyspnoea at diagnosis (n, %)** | 1422 |  |  |  |
| 1-2 |  | 567 (51.2%) | 154 (49.0%) | 0.50 |
| 3-4 |  | 523 (47.2%) | 151 (48.0%) | 0.78 |
| 5 |  | 18 (1.6%) | 9 (2.9%) | 0.16 |
| **GAP stage at diagnosis (n, %)** | 3801 |  |  |  |
| I (points 0-3) |  | 1015 (33.3%) | 532 (70.9%) | **<0.001** |
| II (points 4-5) |  | 1742 (57.1%) | 217 (28.9%) | **<0.001** |
| III (points 6-8) |  | 294 (9.6%) | 1 (0.1%) | **<0.001** |

Males were more likely to have a history of smoking (males n=3229/4432, 72.9% vs females n=718/1187, 60.5%, p<0.001) and have a lower median FVC % predicted at presentation (males 76.4%, IQR 66.2-86.7 vs females 78.8%, IQR 68.6-89.1, p<0.001). There was no statistical difference in the baseline TLCO and KCO based on biological sex (p>0.05).

The list of co-morbidities recorded in the registry is not exhaustive Table 1. Diabetes, cardiovascular disease and gastro-oesophageal reflux disease were all statistically more common in males (p<0.001), whilst major depressive illness was more common in females (p<0.001).

Females were more likely to present with a lower GAP stage compared to males; approximately 70% females had baseline GAP stage I compared to only one third of males (GAP stage 1: males n=1039, 33.8% vs females n=532, 70.9%, p<0.001), although MRC scores did not differ between groups.

### Referral symptoms and diagnosis

At the point of enrolment into the Registry, the majority of patients had experienced symptoms of exertional breathless and/or cough for > 12 months (males n=2584/4246, 60.9% vs females n=736/1171, 62.9%), although significantly more females had experienced symptoms for >24 months (females 40.1% vs males 36.6%, p = 0.028) (Table 2).

**Table 2:** Symptoms and diagnosis of patients with Idiopathic Pulmonary Fibrosis (IPF), stratified by biological sex. Statistically significant values highlighted in bold. Abbreviations: MDT, multidisciplinary team n, number; %, percentage; p value, statistical probability value.

|  | Number of patients with data | Male | Female | P Value |
| --- | --- | --- | --- | --- |
| **Duration of symptoms prior to first appointment** | 5417 |  |  |  |
| No symptoms |  | 85 (2.0%) | 21 (1.8%) | 0.65 |
| Symptoms <6 months |  | 614 (14.5%) | 150 (12.8%) | 0.15 |
| Symptoms 6-12 months |  | 963 (22.7%) | 264 (22.5%) | 0.92 |
| Symptoms 12-24 months |  | 1029 (24.2%) | 266 (22.7%) | 0.28 |
| Symptoms >24 months |  | 1555 (36.6%) | 470 (40.1%) | **0.03** |
| **Diagnosis by consensus MDT (n, %)** | 6019 |  |  |  |
| Discussed at presentation |  | 4394 (93.1%) | 1200 (92.4%) | 0.155 |
| **Radiological pattern (n, %)** | 5117 |  |  |  |
| Definite UIP |  | 1726 (42.8%) | 463 (42.6%) | 0.87 |
| Probable UIP |  | 2081 (51.7%) | 576 (52.9%) | 0.45 |
| Indeterminate |  | 124 (3.1%) | 27 (2.5%) | 0.30 |
| Alternative to UIP |  | 98 (2.4%) | 22 (2.0%) | 0.43 |
| **Surgical biopsy (n,%)** | 5074 |  |  |  |
| Performed |  | 193 ( 4.9%) | 72 (6.6%) | **0.02** |
| Not performed |  | 3774 (95.1%) | 1034 (93.4%) |  |
| **Histological diagnosis (n, %)** |  |  |  |  |
| Definite UIP |  | 114 (2.9%) | 50 (4.5%) | 0.06 |
| Probable UIP |  | 69 (1.7%) | 19 (1.75%) | 0.96 |
| Indeterminate for UIP |  | 10 (0.3%) | 4 (0.4%) | 0.54 |

Most patients with the cohort were discussed in a multidisciplinary team meeting (MDT) to reach a consensus diagnosis (males 93.1% and females 92.4%), with no significant difference across the groups. (Table 2). The presenting radiological pattern of fibrosis on HRCT did not differ statistically between biological sexes, with 94.5% (n=3807/4029) males and 95.5% (n=1039/1088) females having reported definite or probable patterns of UIP (p>0.05).

Whilst the need for a video-assisted thoracoscopic lung biopsy to reach a consensus diagnosis was small across the entire cohort (n=265/5074, 5.2%), more females required a lung biopsy to reach a consensus than did males (females n=72, 6.6%, males n=193, 4.9%, p=0.022, Table 2).

### Management and outcomes

Table 3 provides details on the management of IPF within the registry and correlate with the UK NICE quality standards^17^. Taking the year of presentation into consideration and the baseline FVC % predicted, more males met the FVC NICE eligibility criteria for antifibrotic therapy than females (Pirfenidone: males 54.7% vs females 47.6%, p<0.001 or Nintedanib: males 47.0% vs females 41.5%, p<0.001). A larger proportion of males chose not to start antifibrotic treatment even if they were eligible (males n= 284/604, 47.0% vs females n=118/398, 29.6%, p<0.001). More females were documented within the registry as not meeting eligibility for antifibrotics, despite fulfilling NICE criteria (females n=157/398, 39.4% vs males n=66/604, 10.9%, p<0.001). The reasons for this are not clear. Missing data did not allow comparison of antifibrotic side effect profiles, reasons for stopping antifibrotics or dose reduction between groups.

**Table 3:**
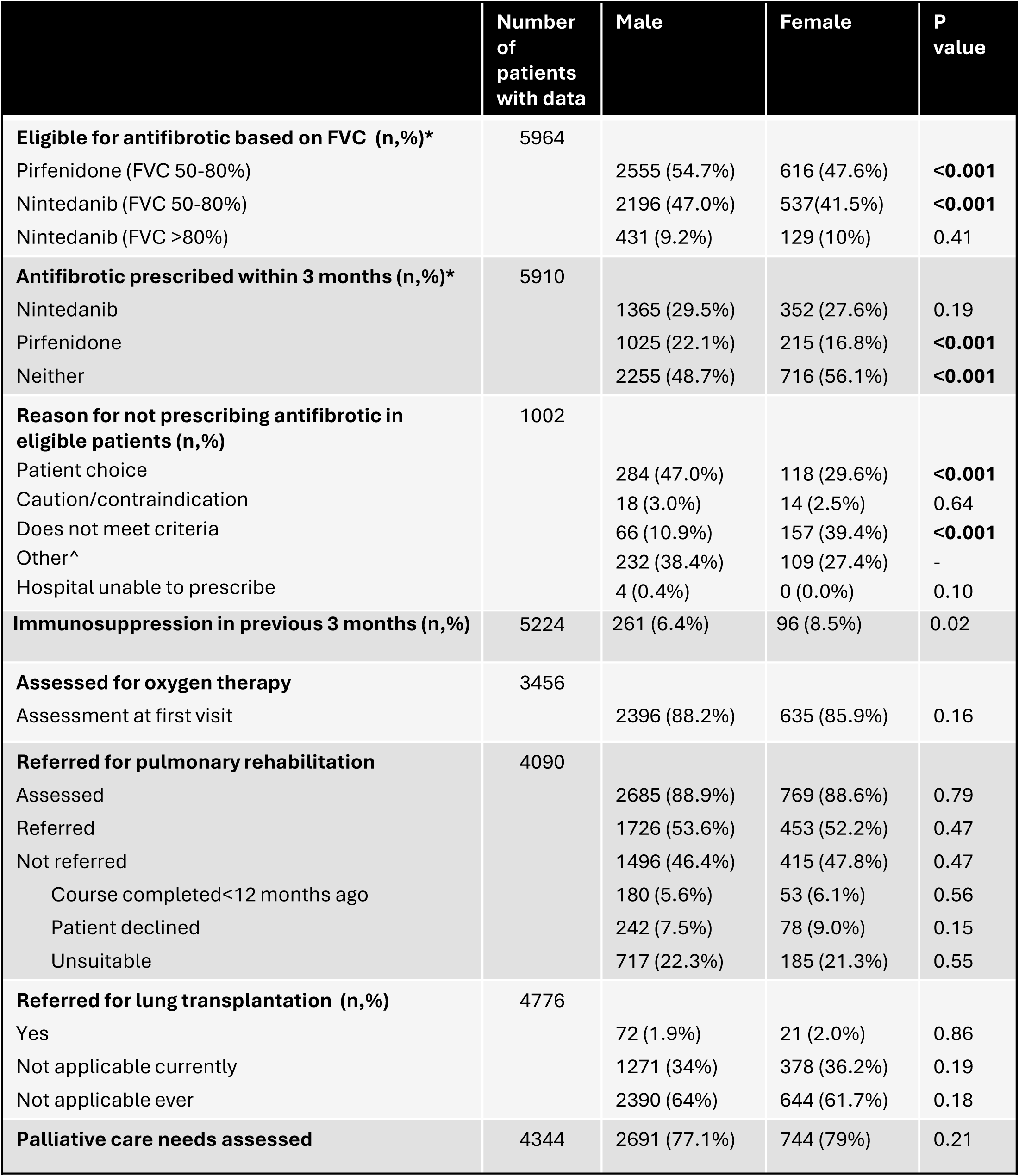
Antifibrotic treatment and management of Idiopathic Pulmonary Fibrosis (IPF) patients at enrolment to registry, stratified by biological sex. Statistically significant values highlighted in bold. Abbreviations: n, number; %, percentage; FVC, forced vital capacity; p value, statistical probability value; ILD, interstitial lung disease; UIP, usual interstitial pneumonia. *subgroups of pirfenidone and nintedanib not mutually exclusive, ^ category not further defined in registry information.

|  | Number<br>of<br>patients<br>with data | Male | Female | P<br>value |
| --- | --- | --- | --- | --- |
| **Eligible for antifibrotic based on FVC (n,%)\*** | 5964 |  |  |  |
| Pirfenidone (FVC 50-80%) |  | 2555 (54.7%) | 616 (47.6%) | **<0.001** |
| Nintedanib (FVC 50-80%) |  | 2196 (47.0%) | 537(41.5%) | **<0.001** |
| Nintedanib (FVC >80%) |  | 431 (9.2%) | 129 (10%) | 0.41 |
| **Antifibrotic prescribed within 3 months (n,%)\*** | 5910 |  |  |  |
| Nintedanib |  | 1365 (29.5%) | 352 (27.6%) | 0.19 |
| Pirfenidone |  | 1025 (22.1%) | 215 (16.8%) | **<0.001** |
| Neither |  | 2255 (48.7%) | 716 (56.1%) | **<0.001** |
| **Reason for not prescribing antifibrotic in eligible patients (n,%)** | 1002 |  |  |  |
| Patient choice |  | 284 (47.0%) | 118 (29.6%) | **<0.001** |
| Caution/contraindication |  | 18 (3.0%) | 14 (2.5%) | 0.64 |
| Does not meet criteria |  | 66 (10.9%) | 157 (39.4%) | **<0.001** |
| Other^ |  | 232 (38.4%) | 109 (27.4%) | - |
| Hospital unable to prescribe |  | 4 (0.4%) | 0 (0.0%) | 0.10 |
| **Immunosuppression in previous 3 months (n,%)** | 5224 | 261 (6.4%) | 96 (8.5%) | 0.02 |
| **Assessed for oxygen therapy** | 3456 |  |  |  |
| Assessment at first visit |  | 2396 (88.2%) | 635 (85.9%) | 0.16 |
| **Referred for pulmonary rehabilitation** | 4090 |  |  |  |
| Assessed |  | 2685 (88.9%) | 769 (88.6%) | 0.79 |
| Referred |  | 1726 (53.6%) | 453 (52.2%) | 0.47 |
| Not referred |  | 1496 (46.4%) | 415 (47.8%) | 0.47 |
| Course completed<12 months ago |  | 180 (5.6%) | 53 (6.1%) | 0.56 |
| Patient declined |  | 242 (7.5%) | 78 (9.0%) | 0.15 |
| Unsuitable |  | 717 (22.3%) | 185 (21.3%) | 0.55 |
| **Referred for lung transplantation (n,%)** | 4776 |  |  |  |
| Yes |  | 72 (1.9%) | 21 (2.0%) | 0.86 |
| Not applicable currently |  | 1271 (34%) | 378 (36.2%) | 0.19 |
| Not applicable ever |  | 2390 (64%) | 644 (61.7%) | 0.18 |
| **Palliative care needs assessed** | 4344 | 2691 (77.1%) | 744 (79%) | 0.21 |

Although not a recommended treatment for IPF (NICE CG163)^18^, females were statistically more likely to have received immunosuppressive medications during their treatment pathway (females 8.5% vs males 6.4%, p=0.015). There was no significant difference in percentages of patients assessed for oxygen, pulmonary rehabilitation or lung transplantation, based on biological sex.

Female sex was associated with longer mean survival (females 9.99 years (95% CI 9.18-10.79) vs males 8.57 years (95% CI 8.15-8.99), p<0.001) (Figure 1). Cox regression was performed on a cohort of 713 patients with complete datasets for FVC % predicted, age, sex, smoking and co-morbidities with at least 5 years follow up (patients registered up to and including October 2019). Male sex, higher age, lower baseline FVC % predicted and co-existent lung cancer were all independently associated with worse survival (Figure 1). The strongest independent association with IPF survival was co-existent lung cancer (HR 9.3, 95% C.I. 2.86-30.24, p<0.001) whilst male sex was associated with HR of 1.76 (95% C.I. 1.22-2.54, p<0.002). Interaction terms indicated that there was no significant difference n the effect of age, FVC, smoking status or comorbidities on survival between sexes (p>0.05, data not shown).

**Figure 1.**
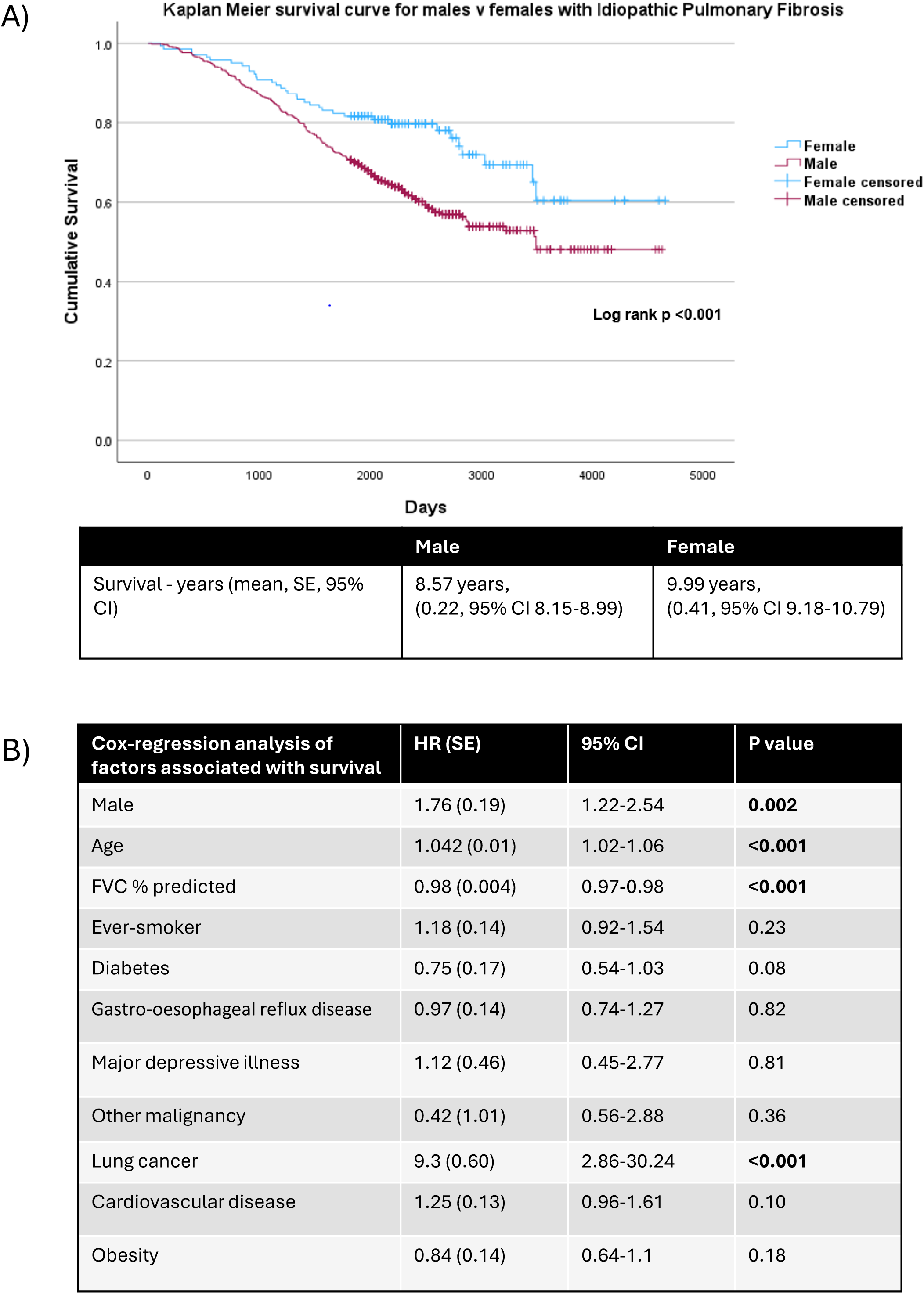
A) Kaplan Meier survival curve for Idiopathic Pulmonary Fibrosis (IPF) patients stratified by biological sex. Female sex was associated with longer mean survival (females 9.99 years (95% CI 9.18-10.79) vs males 8.57 years (95% CI 8.15-8.99), p<0.001). B) Cox-regression analysis of factors independently associated with survival. Statistically significant values highlighted in bold. Abbreviations: HR, Hazards ratio; 95%CI, 95% confidence interval; P value, statistical probability value; %, percentage; SE, standard error

## Discussion

This is the first UK study to use BTS-ILD registry data to evaluate the baseline characteristics, treatment, management and outcomes of patients with IPF, stratifying by biological sex. The results of this study suggest that IPF has distinct characteristics depending on biological sex with notable differences in baseline disease severity, co-morbidities, duration of symptoms prior to referral, need for lung biopsy to make a consensus MDT diagnosis and overall survival. The study highlights key areas that should be explored to understand potential reasons underlying these sex differences in IPF, to facilitate tailored based care and to improve patient outcomes.

In this cohort males presented with more severe disease, having a lower FVC % predicted and higher GAP stage than females at baseline. This is in keeping with the findings of a multicentre prospective French IPF cohort (n= 236, COhorte FIbrose, COFI)^5^, who found that females tended to present with less advanced disease at diagnosis, better lung function, and more preserved FVC. The authors hypothesised this might be due to lower exposure to triggering agents such as tobacco smoke or occupational inhalants compared to men, although recognised future changes in lifestyle habits may influence this. Reflecting on the differing smoking history exposure in our own cohort, our findings could support this hypothesis.

Barriers to timely diagnosis in ILD are well described^19^ and the UK BTS ILD registry data has previously highlighted the negative impact that even 6-12 months delay in assessment can have on survival in IPF^20^. The data from this cohort is no exception. Patients experienced a significant length of time between the onset of their symptoms and clinical assessment.

Furthermore, more females had symptoms for >24 months at their first clinic visit compared to males. Whilst some of these observations may be related to workforce related challenges and waiting times, it may also relate to differences in lived experience of the disease. Han et al demonstrated that independent of age, disease severity or functional capacity, females have greater mental health impairment but fewer physical health related QoL impairments than males^2^. The associations between self-efficacy, social support and sex pattern in PF are unknown. Gendered expectations may be an influencing factor and worthy of further exploration.

Sex differences were identified in the frequency of the dominant co-morbidities. Although the list recorded in the registry was not exhaustive, cardiovascular disease, gastro-oesophageal reflux disease and diabetes were more common in males, whilst major depressive illness was more common in females. This is in keeping with a study of 348 Swedish IPF registry participants, where coronary artery disease and other cardiovascular diseases, such as atrial fibrillation and heart failure were more prevalent in males, but thyroid disease and osteoporosis were more common in females^3^. Whilst shared risk factors e.g. smoking history, may explain this finding, Izbicki et al. ^21^ found that the association between IPF and increased risk of developing coronary artery disease was independent of these common confounding factors, but a mechanistic explanation for this observed association was not clearly defined.

Previous studies have indicated that definite or probable UIP patterns of fibrosis on HRCT are more commonly found in male patients, facilitating an IPF diagnosis. Conversely, atypical radiological patterns are more frequent in female patients, necessitating a surgical lung biopsy to confirm the diagnosis of IPF^5, 22^. In this cohort we found that whilst there was no significant difference in the proportions of HRCTs demonstrating definite or probable UIP patterns, more females required a lung biopsy to secure an MDT diagnosis of IPF. Assayag et al. have previously investigated the role of gender in making a confident diagnosis of IPF and concluded there is evidence for bias when using sex to inform an ILD diagnosis, which may result in men being over-diagnosed and women underdiagnosed with IPF^22, 23^. The finding that statistically more females were prescribed immunosuppressive medications, not a currently recommended treatment strategy for IPF (CG163), may suggest a degree of unconscious bias towards non-IPF diagnoses of these patients during their pathway.

The data suggests that whilst males were more likely to meet NICE eligibility criteria for antifibrotics at baseline, there was a significant proportion of males that declined to take antifibrotics through patient choice, and this was significantly higher than in females (males 47.0% vs females 29.6%). Other real-world datasets have suggested that both sex and ethnicity may affect uptake of antifibrotic medications, with the potential for subsequent impact on patient outcomes. A study of n=14,792 US veterans with IPF showed that whilst uptake of antifibrotics was generally low in this cohort (17% overall), there were significant disparities in adoption, with lower uptake associated with female sex (adjusted OR, 0.41; 95% CI, 0.27-0.63; p<0.001), Black race (adjusted OR, 0.60; 95% CI, 0.49-0.73; p<0.001), and rural residence (adjusted OR, 0.88; 95% CI, 0.80-0.97; p= 0.012)^24^. Similarly, Ghimire et al. 2024 demonstrated racial and sex disparities in antifibrotic prescriptions in a large US cohort of IPF patients (n=10,667)^25^. Although disease prevalence was relatively even amongst sexes (46.6% males vs 53.3% females), there was a significantly higher prescription of antifibrotics amongst Caucasians and males (76.6% males vs 39.4% females), with improved hospital outcomes for those on antifibrotic therapy (information on lung function testing was not provided).

In this cohort there were insufficient complete datasets available to compare antifibrotic side effect profiles, reasons for stopping antifibrotics or dose reduction between groups. Adherence to antifibrotic treatment in IPF was recently studied in an Italian prospective cohort. Among 667 new IPF cases, 296 received antifibrotic prescriptions (77% male), with 62.8% being adherent in the first year. Whilst the number of female patients receiving antifibrotics was lower, adherence was not significantly associated with sex or age, co-morbidity score, the number of concomitant drugs, or antifibrotic used^26^. Conversely, Ortiz et al.^27^ showed in their multicentre Spanish cohort of 232 IPF patients commencing antifibrotics, that whilst time to first adverse event and all-cause mortality were similar between the antifibrotic medications, females were more likely to withdraw from therapy due to side effects (predominantly gastrointestinal). Female sex, diarrhoea and photosensitivity were independent factors associated with an increased risk for definitive discontinuation from therapy. Furthermore, post-hoc analyses of clinical trial data have shown more frequent dose reductions, treatment interruptions and early discontinuations of antifibrotic medications in females^28^. Given that females were significantly underrepresented in the clinical trials of antifibrotics, additional real-world studies are required to explore these factors in more detail.

In this UK cohort, females demonstrated a significantly better survival outcome compared to males, with worse survival associated independently with male gender, increasing age, lower baseline FVC, and co-existent lung cancer. This is in keeping with the large body of evidence that contributed to the development of the GAP (Gender, age and physiology) model used to help predict mortality in IPF^16^. In a single centre retrospective study of IPF patients in Italy^29^, women also demonstrated better survival and better tolerance to long-term therapy compared to males. Interestingly, no gender differences emerged in terms of reduction in/discontinuation of therapy, nor did this impact mortality. There are however contrasting reports as to the influence of sex and gender on survival in the broader literature. A multicentre, prospective study of French IPF patients demonstrated no survival differences based on biological sex^5^, although the authors comment that this may have been due to small sample size and lack of statistical power. The UK parliamentary brief on Men’s Health, gives important context to the significant disparities in male life expectancy, between some groups of men within the UK and across a range of diseases^30^. The reasons underpinning this are likely to be numerous and have interconnected influence; health engagement, health literacy, social determinants and behaviour can all vary with an individual’s culture, community and soci-economic status. Consequently, several stakeholders and experts have called for a national men’s health strategy in the UK.

As with all real-world data sets there are limitations. The first limitation concerns that of missing data which we have reflected transparently upon within the results section. We acknowledge that the statistical robustness of the survival data and generalisation of the cox-regression results to the entire cohort, may have been biased by both missing data and the immaturity of data in terms of follow-up. Nonsensical data entry also contributed to missing data items, for example input of oxygen saturation of 0%. A recent update to the registry platform (November 2024) has ensured that it is now impossible to enter data that falls outside strict validation boundaries and should improve the quality of data received. Secondly, data entry for specialist centres was mandated in 2022 but was voluntary prior to that date and remains so for participating secondary care hospitals. Data entry is therefore dependent on local resource allocation, which was significantly impacted during the COVID pandemic, resulting in low data entry during 2020-2021. Mandated data entry for specialist centres will hopefully drive future patient enrolment. Finally, most patients were enrolled by specialist centres and thus the dataset may not be generalisable to all IPF patients seen within secondary care.

## Conclusion

The results of this study suggest that IPF has distinct characteristics depending on biological sex. Additional research is required to explore and understand potential reasons underlying sex differences in IPF to facilitate tailored based care and to improve patient outcomes. The recorded data did not allow for analysis by gender. Gender, separate to biological sex, may account for, or contribute to, differences noted in the data and this should be explored so that it can be accounted for, or absented accordingly. In order to achieve this it is important that future clinical trials explicitly explore sex and gender-specific targeted interventions and undertake detailed gender and sex analyses, taking into account different presentations, environmental risk factors and lived experiences of this population to optimise future IPF patient care.

## Data Availability

All data produced in the present work are contained in the manuscript

## Contributors

SM and SB were responsible for the initial draft of the paper and SB is the guarantor. PW and GD provided statistical support. AMR, SH, MW provided additional revisions. All authors reviewed and approved the final manuscript.

## Acknowledgements

We would like to acknowledge all participating hospitals as in Appendix 1 for their time and dedication in inputting the data into the registry template and the patients who gave consent for their anonymised data to be used. We would also like to thank the Bristish Thoracic Society (BTS) and BTS ILD steering committee for their support in publication of this article.

## Disclaimer and data availability statement

This publication makes use of data purchased from the British Thoracic Society Interstitial Lung Disease Registry Programme, which has no responsibility or liability for the accuracy, currency or correctness of this publication. The project was peer reviewed by the BTS prior to release of the data.

## Statement of Ethics

The UK ILD (Interstitial lung disease) registry has ethical approval (National Research Ethics Service (NRES) reference 12/EE/0381 and renewal 12/EE/0346). A grant from the Healthcare Quality Improvement Partnership (HQIP) (2012-2014) supported the initial development of the Registry, whilst financial support from Boehringer Ingelheim and Intermune also helped to augment the data collection software in 2014.

## References

1. Raghu G, Weycker D, Edelsberg J, Bradford WZ, Oster G. Incidence and prevalence of idiopathic pulmonary fibrosis. Am J Respir Crit Care Med. 2006;174(7):810–6.

2. Han MK, Swigris J, Liu L, Bartholmai B, Murray S, Giardino N, et al. Gender influences Health-Related Quality of Life in IPF. Respir Med. 2010;104(5):724–30.

3. Kalafatis D, Gao J, Pesonen I, Carlson L, Skold CM, Ferrara G. Gender differences at presentation of idiopathic pulmonary fibrosis in Sweden. BMC Pulm Med. 2019;19(1):222.

4. Zaman T, Moua T, Vittinghoff E, Ryu JH, Collard HR, Lee JS. Differences in Clinical Characteristics and Outcomes Between Men and Women With Idiopathic Pulmonary Fibrosis: A Multicenter Retrospective Cohort Study. Chest. 2020;158(1):245–51.

5. Sese L, Nunes H, Cottin V, Israel-Biet D, Crestani B, Guillot-Dudoret S, et al. Gender Differences in Idiopathic Pulmonary Fibrosis: Are Men and Women Equal? Front Med (Lausanne). 2021;8:713698.

6. Redente EF, Jacobsen KM, Solomon JJ, Lara AR, Faubel S, Keith RC, et al. Age and sex dimorphisms contribute to the severity of bleomycin-induced lung injury and fibrosis. Am J Physiol Lung Cell Mol Physiol. 2011;301(4):L510–8.

7. Costabel U, Inoue Y, Richeldi L, Collard HR, Tschoepe I, Stowasser S, et al. Efficacy of Nintedanib in Idiopathic Pulmonary Fibrosis across Prespecified Subgroups in INPULSIS. Am J Respir Crit Care Med. 2016;193(2):178–85.

8. Vancheri C, Sebastiani A, Tomassetti S, Pesci A, Rogliani P, Tavanti L, et al. Pirfenidone in real life: A retrospective observational multicentre study in Italian patients with idiopathic pulmonary fibrosis. Respir Med. 2019;156:78–84.

9. Assayag D, Garlick K, Johannson KA, Fell CD, Kolb M, Cox G, et al. Treatment Initiation in Patients with Interstitial Lung Disease in Canada. Ann Am Thorac Soc. 2021;18(10):1661–8.

10. Spencer LG, Loughenbury M, Chaudhuri N, Spiteri M, Parfrey H. Idiopathic pulmonary fibrosis in the UK: analysis of the British Thoracic Society electronic registry between 2013 and 2019. ERJ Open Res. 2021;7(1).

11. Fahim A, Loughenbury M, Stewart I, Agnew S, Almond H, Casimo L, et al. Idiopathic pulmonary fibrosis in the UK: findings from the British Thoracic Society UK Idiopathic Pulmonary Fibrosis Registry. BMJ Open Respir Res. 2025;12(1).

12. Raghu G, Collard HR, Egan JJ, Martinez FJ, Behr J, Brown KK, et al. An official ATS/ERS/JRS/ALAT statement: idiopathic pulmonary fibrosis: evidence-based guidelines for diagnosis and management. Am J Respir Crit Care Med. 2011;183(6):788–824.

13. TA282 NICE-TAG. Pirfenidone for treating idiopathic pulmonary fibrosis https://www.nice.org.uk/Guidance/TA2822013 [Published 2013, Accessed 4th March 2025].

14. TA379 NICE-TAG. Nintedanib for treating Idiopathic Pulmonary Fibrosis https://www.nice.org.uk/guidance/ta3792016 [Published 2016, Accessed 3rd March 2025].

15. TA864 NICE-TAG. Nintedanib for treating idiopathic pulmonary fibrosis when forced vital capacity is above 80% predicted https://www.nice.org.uk/guidance/ta8642023 [Published 2023, Accessed 28th Feb 2025].

16. Ley B, Ryerson CJ, Vittinghoff E, Ryu JH, Tomassetti S, Lee JS, et al. A multidimensional index and staging system for idiopathic pulmonary fibrosis. Ann Intern Med. 2012;156(10):684–91.

17. QS79 NICE-Quality Standard. Idiopathic pulmonary fibrosis in adults https://www.nice.org.uk/guidance/qs792015 [Published 2015, Accessed 3rd March 2025].

18. CG163 NICE-Clinical Guideline. Idiopathic Pulmonary Fibrosis in adults: diagnosis and management https://www.nice.org.uk/guidance/cg163: National Institure for Health Care Excellence [Published 2013, Updated 2017, Accessed 3rd March 2025].

19. Cosgrove GP, Bianchi P, Danese S, Lederer DJ. Barriers to timely diagnosis of interstitial lung disease in the real world: the INTENSITY survey. BMC Pulm Med. 2018;18(1):9.

20. Shankar R, Hadinnapola CM, Clark AB, Adamali H, Chaudhuri N, Spencer LG, et al. Assessment of the impact of social deprivation, distance to hospital and time to diagnosis on survival in idiopathic pulmonary fibrosis. Respir Med. 2024;227:107612.

21. Izbicki G, Ben-Dor I, Shitrit D, Bendayan D, Aldrich TK, Kornowski R, et al. The prevalence of coronary artery disease in end-stage pulmonary disease: is pulmonary fibrosis a risk factor? Respir Med. 2009;103(9):1346–9.

22. Assayag D, Morisset J, Johannson KA, Wells AU, Walsh SLF. Patient gender bias on the diagnosis of idiopathic pulmonary fibrosis. Thorax. 2020;75(5):407–12.

23. Strek ME. Gender in idiopathic pulmonary fibrosis diagnosis: time to address unconscious bias. Thorax. 2020;75(5):365–6.

24. Kaul B, Lee JS, Petersen LA, McCulloch C, Rosas IO, Bandi VD, et al. Disparities in Antifibrotic Medication Utilization Among Veterans With Idiopathic Pulmonary Fibrosis. Chest. 2023;164(2):441–9.

25. Ghimire P. GS, Thameem D. . Racial and gender disparities, and impact of antifibrotic therapy on hospital outcomes in patients with Idiopathic Pulmonary Fibrosis [Abstract]. Chest. 2024;166(4):A3193.

26. Iommi M, Gonnelli F, Bonifazi M, Faragalli A, Mei F, Pompili M, et al. Understanding Patterns of Adherence to Antifibrotic Treatment in Idiopathic Pulmonary Fibrosis: Insights from an Italian Prospective Cohort Study. J Clin Med. 2024;13(9).

27. Romero Ortiz AD, Jimenez-Rodriguez BM, Lopez-Ramirez C, Lopez-Bauza A, Perez-Morales M, Delgado-Torralbo JA, et al. Antifibrotic treatment adherence, efficacy and outcomes for patients with idiopathic pulmonary fibrosis in Spain: a real-world evidence study. BMJ Open Respir Res. 2024;11(1).

28. Jouneau S, Gamez AS, Traclet J, Nunes H, Marchand-Adam S, Kessler R, et al. A 2-Year Observational Study in Patients Suffering from Idiopathic Pulmonary Fibrosis and Treated with Pirfenidone: A French Ancillary Study of PASSPORT. Respiration. 2019;98(1):19–28.

29. Tondo P, Scioscia G, De Pace CC, Murgolo F, Maci F, Stella GM, et al. Gender Differences Are a Leading Factor in 5-Year Survival of Patients with Idiopathic Pulmonary Fibrosis over Antifibrotic Therapy Reduction. Life (Basel). 2025;15(1).

30. Ramsey DA. BS. Parliamentary Office of Science and Technology (POST). POSTbrief 56: Men’s Health. UK Parliament https://researchbriefings.files.parliament.uk/documents/POST-PB-0056/POST-PB-0056.pdf [Published 2023, Accessed 11th March 2025.

